# The relative power of individual distancing efforts and public policies to curb the COVID-19 epidemics^*^

**DOI:** 10.1101/2020.07.06.20147033

**Authors:** Cécile Aubert, Emmanuelle Augeraud-Véron

## Abstract

Lockdown curbs the COVID-19 epidemics but at huge costs. Public debates question its impact compared with reliance on individual responsibility. We aim at understanding how rationally chosen self-protective behavior impacts the spread of the epidemics. We want to, first, assess the value of lockdown compared to a counterfactual that incorporates self-protection efforts under unknown disease prevalence; and second, assess how individual behavior modify the epidemic dynamics when mandatory policies are relaxed. We couple an SLIAR model, that includes asymptomatic transmission, with utility maximization: Individuals trade off economic and wellbeing costs from physical distancing with a lower infection risk. Effort depends on risk aversion, perceptions, and the value of contacts. In a Nash equilibrium, individual uncoordinated efforts yield average contact intensity, which drives epidemic transmission. Equilibrium effort differs markedly from constant, stochastic or proportional contacts reduction. It adjusts to reported cases in a way that creates a slightly decreasing plateau in epidemic prevalence. Calibration on French data shows that the number of deaths with no lockdown but equilibrium efforts is only 1/6 to 1/10 of the number predicted with business-as-usual. However, lockdown saves at least 50% more lives than individual efforts alone. Prolonged weaker restrictions prevent an exponential rebound. Public policies post-lockdown have a limited impact as they partly crowd out individual efforts. Compulsory mask wearing helps resume activity but has no impact on the epidemic. Communication that increases risk salience is more effective.

## Introduction

The COVID-19 pandemic is largely controllable with non-pharmaceutical interventions such as physical distancing, especially through lockdown. The latter has curbed the spread of the epidemics during winter 2020 in Wuhan and Shanghai [1, 2], where contacts have been reduced 7-to 8-fold [3]. But such a drastic reduction involves huge economic and welfare costs [4]. As of June 21, 2020, the COVID-19 pandemic has caused more than 468,000 deaths worldwide [5] (US: 120,000, Brazil: 50,000, UK: 42,000) and huge economic losses. Relaxing lockdown allows limiting economic disruption. Yet some countries had to reintroduce constraints after lifting lockdown (Germany and South Korea in May, Germany and China in June). With a rising tension between sanitary and economic impacts, political debates question whether lockdown is necessary or excessive, since individuals would take precautions if left to choose their behavior.

Scientific evidence about the benefits of lockdown does not account for individuals’ efforts. [6] estimate that lockdown has saved 3 million lives in 11 European countries up to May 4, 2020, using constant *R*_0_ values that change with policy interventions. [7] estimate that lockdown-related measures have avoided 500 million cases in in China, South Korea, Italy, Iran, France and the United States between January and April 6, 2020. However, [8] use Google mobility data and find that individuals largely adjusted their behavior before restrictions were imposed. Google’s work related mobility index negatively correlates with virus reported prevalence even in the absence of compulsory measures.

To estimate the ‘net impact’ of lockdown, we build a counterfactual in which transmission rates are driven by individual rational choices. We couple an SLIAR compartmental model (with asymptomatic and symptomatic infectious individuals [9]), and a utility maximization model of self-protection under risk. The epidemic model differs from standard ones [10, 11] to reflect the high proportion of infectious individuals with no or only mild symptoms [12]. Asymptomatic transmission is key in determining the effectiveness of public policies [13]. It also makes it difficult to estimate the infection risk. Individuals base their perception on the most salient information available in the media, the number of reported cases. Distancing decisions are the outcome of a Nash equilibrium: Going out is more or less risky depending on others’ choice to go out. The epidemic transmission rate depends on equilibrium distancing efforts; and thus indirectly on individuals’ risk aversion, costs to avoiding contacts, and beliefs.

Epidemiological parameters are fitted on French data. The start of lockdown is very well identified in France (contrary to countries where is was more gradual). French citizens could use information from Italy to better adapt their behavior, since the epidemic pattern was similar, with a 7-day lag. We compute the number of deaths one would have experienced in the absence of lockdown but with individual self-protection efforts, and compare it to simulations based on business as usual and to the actual number of deaths under lockdown. Our results contradict both the large estimates based on business-as-usual and the idea that individual efforts would be as effective as lockdown. We also identify general results about the impact of alternative policies after the end of global lockdown, such as wearing a mask. Public policies that are effective at reducing reported severe cases, induce a reduction in equilibrium efforts. This countervailing effect very largely undermines their efficiency.

## Materials and methods

### Data

We use publicly available data from the French national health agency, covering the period between February 25 and June 17, 2020. The starting date of the lockdown is clearly identified at March 17, 2020: Different policies pertaining to lockdown were taken nearly at the same time (school closure, closure of non-essential business facilities, confinement to a 1-kilometer range from home, outside leisure activities for no more than 1 hour per day…). Data corresponds to different policy stages, for which we use different fitting methods: pre-lockdown ([*t*_0_, *t*_1_], 02/25 – 03/17), full lockdown ([*t*_1_, *t*_2_], 03/17 – 05/11), intermediate lockdown ([*t*_2_, *t*_3_], 05/11 – 06/17). Data reporting has been somewhat irregular but the counting method has been consistent until June 3. These data are freely available at https://dashboard.covid19.data.gouv.fr/vue-d-ensemble?location=FRA.

### The epidemiological model

We integrate in an SLIAR compartmental epidemiological model [9, 14–16] an economic model of self-protection choices [17–20]. The SLIAR model allows accounting for asymptomatic or mild symptomatic transmission. A specific feature of COVID-19 is indeed the high proportion of asymptomatic but infectious individuals. The population is separated into *susceptible* individuals (in number *S*(*t*) at date *t*) who can get contaminated, infectious *latent* individuals (*L*(*t*)), *asymptomatic* infectious individuals (*A*(*t*)), who are not reported as being ill, and symptomatic *infectious* individuals (*I*(*t*)) who know that they have the disease. Symptomatic infectious individuals are the only ones counted in reported cases. They may have been tested positive or may have severe, unmistakable, symptoms. They are isolated or hospitalized, and no longer play a significant part in disease transmission. Contamination is due to latent and asymptomatic infectious individuals (in number *L*(*t*) + *A*(*t*)).

At the population level, the dynamics is given by:

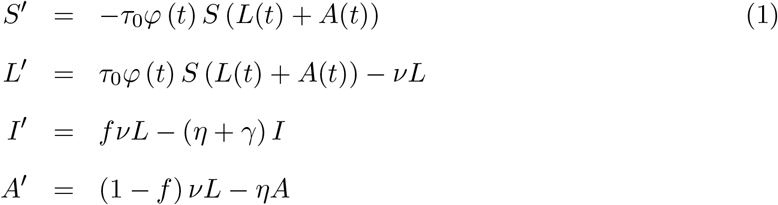

Contaminated susceptible individuals become latent infectious for an average time *ν*^*−*1^. Then, a fraction *f* of latent infectious individuals develop severe symptoms, which last for a mean time *η*^*−*1^ after which they are removed and fully immunized. A fraction 1 *− f* of latent infectious individuals become mild symptomatic and lose infectiousness after an average time *η*^*−*1^. They then recover and are immunized and removed from disease transmission. However, severe symptomatic individuals may also die at rate *γ*. Note that as the recovered play no role in epidemics transmission nor in risk perception, we only need to describe the dynamics for the susceptible and the three infectious compartments.

A specificity of our analysis is that the transmission rate *τ*_0_*φ* (*t*) depends on the infectiousness of the disease but also on individual physical distancing decisions. We denote *τ*_0_ the transmission rate of the disease that would apply in the absence of individual behavior adaptation to the spread of the disease (‘business-as-usual’). This is the transmission rate that can be observed at the beginning of the epidemics in the region considered. Transmission rate at time *t* is the product of *τ*_0_ times contact intensity *φ*(*t*), defined as the ratio of the number of contacts at time *t* over the number of contacts at the beginning of the epidemic outbreak. This contact intensity models physical distancing.

The aim of this work is to propose modelling of the contact intensity function under various scenarios and to compare the spread of the epidemics according to these scenarios.

### The behavioral model

In the absence of legal constraints, contact intensity *φ*(*t*) is set by the self-protection effort *ε* that individuals choose. Self-protection effort *ε* is the outcome of a Nash equilibrium, in which individuals choose a best response to their economic, psychological and epidemiological environment. [21] and [22] also endogenize transmission to economic incentives, in different and convincing ways, but they consider a classical SIR model with infectious individuals who are aware of their immune status. We incorporate asymptomatic transmission and various preference determinants. [23] and [24] had already highlighted that while epidemic-driven transmission increases in the number of infected individuals, economically-driven transmission decreases with this number, due to higher effort. We study how the two dynamics interact.

### Perceived infection risk

To determine self-protection efforts *ε*, we need to specify how the infection risk is perceived. The perceived infection risk function 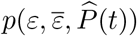, depends on personal exposure (the self-protection effort *ε*), on the average physical distancing effort 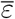 in the population and on perceived prevalence 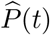. We assume the following specification.

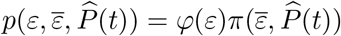

Function *φ* (*ε*) *∈* |0, 1] describes the contact intensity resulting in doing a self-protection effort *ε*. Thus, decreasing function *φ* (*ε*) describes the reduction in risk due to more individual effort. Perceived disease prevalence, 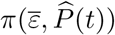, decreases in others’ average behavior 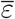 and is an increasing function of perceived prevalence 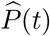.

Perceived prevalence 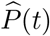 is a function of the salient information available in the media [25], that is, daily reported cases *fνL*. Individuals use information that is not complete (the number of individuals involved in the disease transmission is not known). They may however not underestimate the infection risk as it is very salient. Perceived prevalence incorporates a weight *k, k >* 1, that reflects extra attention paid to the COVID-19 risk, awareness that many infectious individuals are undetectable, and over-weighing of small probabilities in self-protection decisions [26]. Thus

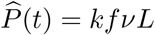

Thus perceived infection risk would be denoted 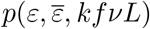.

### Individual distancing efforts and the corresponding Nash equilibrium

Each individual independently chooses an effort *ε* to reduce her contacts, and thereby reduce her perceived probability of infection at each date. However, reducing contacts creates a disutility, as it involves psychological costs and economic foregone opportunities (like working only very few hours). The chosen self-protection effort *ε* maximizes at time *t* the individual’s expected utility defined as:

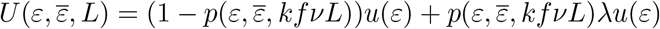

Function *u*(*ε*) is the von Neumann and Morgenstern utility function. It is concave to represent risk aversion and decreasing in distancing effort *ε*. Because reducing contacts affects economic opportunities and wellbeing when sane, the associated cost is part of the utility function, and is affected by risk aversion. To allow for calibrations, we consider a standard power utility function: 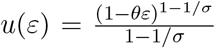, where *θ* measures the utility (economic and welfare) loss due to contacts reduction and *σ* is the constant relative risk aversion (CRRA) parameter. Parameter *λ* measures the loss in utility when becoming sick.

The impact of reducing own social contacts depends on the average effort undertaken in the population 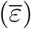. In large number population, each individual does not take into account how her contacts reduction efforts affects the infection risk of others. Thus, the optimal individual effort *ε* is the best response function

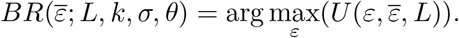

The Nash equilibrium physical distancing effort *ε*^*\**^ is determined as a fixed point of best response function *BR* and solves

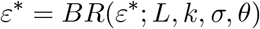

We denote the corresponding contact intensity *φ*(*ε*^*\**^(*L*(*t*))) = *φ*^*\**^(*L*(*t*)).

## Results

We characterize the equilibrium distancing effort in the absence of lockdown, and use it to contrast the number of deaths under three cases: full lockdown (data), ‘business-as-usual’, and equilibrium effort (counterfactual). We then assess the impact of the prolonged intermediate lockdown phase on epidemic dynamics given individual choices.

### Equilibrium physical distancing effort

We assume a linear impact of effort *φ*(*ε*) = 1 *− ε ∈* [0, 1] and 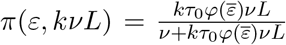. The individual physical distancing effort is computed as 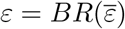, with

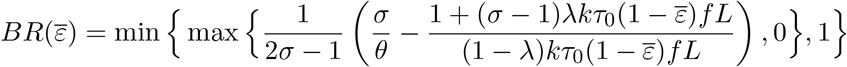

The symmetric Nash equilibrium is the solution to equation *ε*^*\**^ = *BR*(*ε*^*\**^), under the constraints that this solution lies in [0,1]. It can be explicitly computed as

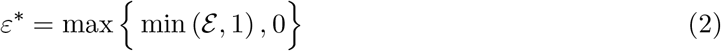

where

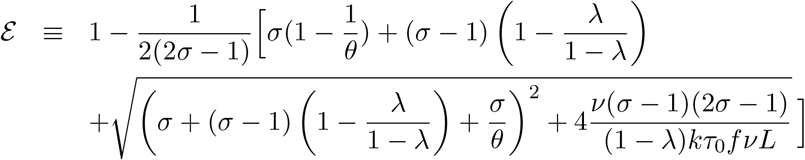

All parameters change the equilibrium physical distancing effort in the expected direction. Their impact is complex and quite different from proportionality. Equilibrium effort increases non linearly in *risk aversion σ*, and in *perceptions* about disease severity *λ* and about prevalence *k*. It decreases non linearly in *personal economic and welfare costs θ*.

### The impact of lockdown on number of deaths

We study the epidemic dynamics under various scenarios on contact intensity during lockdown [*t*_1_, *t*_2_]. The list of epidemiological parameters is given in S1-Table. The counterfactual situations we consider enable to better understand the net impact of lockdown on the number of deaths.

### Counterfactual with equilibrium efforts

Our characterization of equilibrium effort is used to simulate the counterfactual epidemic dynamics if there had been no lockdown, but individual self-protection. The epidemiological dynamics are given by System 1, with counterfactual contact intensity function *φ*(*t*) = *φ*_*CF*_ (*t*) defined as

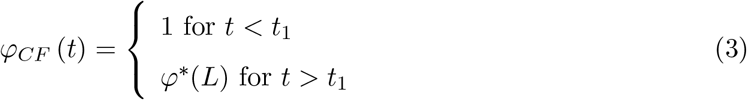

Parameters in the utility function are set at *θ* = 0.1, *λ* = 0.1, and the CRRA coefficient of risk aversion, *σ*, at *σ* = 1.5 [27].

### Calibration of lockdown on the data

We compare the counterfactual dynamics above to simulations based on business-as-usual (no reduction in contacts intensity, i.e., *φ* = 1) and to lockdown, which is fitted on the data.

The lockdown situation has been calibrated in [28]. The impact of lockdown on contact intensity is modeled with a time dependent function *φ*_*L*_, defined by System 4. Function *φ*_*L*_ is set to 1 for the period before public health measures had been taken ([*t*_0_, *t*_1_]), up to *t*_1_ = March 17). The impact of these interventions is not instantaneous [29]. An exponential form can model this delay, as in [30] for the 2014 Ebola virus outbreak or [15] for COVID-19 early stages. But because the French lockdown was very long (9 weeks in its strictest form), we use a form that has a better fit, similar to [29], where *φ*_*L*_(*t*) decreases from 1 to *a >* 0. Function *φ*_*L*_(*t*) is given by Eq. (4).

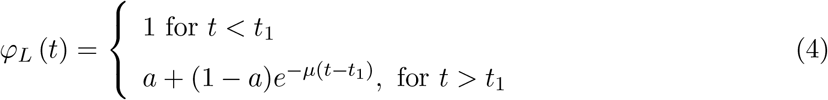

Parameters *a, µ* have been calibrated with least-squares fitting. We also use the lockdown simulation and the data to fit mortality parameter *γ*. The calibration resulted in: *a* = 0.1828483, *µ* = 0.1424125 and *γ* = 0.0203774. Thus, contacts have been reduced 5-6 fold in France during lockdown (compared to 7-8 fold in Wuhan and Shanghai [31], which is consistent since the French lockdown was less strict).

### Contact intensity under lockdown vs. under equilibrium effort

Fig. 1 represents contact intensity, and corresponding effort, under lockdown and in the rational effort equilibrium (given simulated epidemic prevalence, 1). Business-as-usual corresponds to a contact intensity equal to 1 by definition.

**Figure 1:**
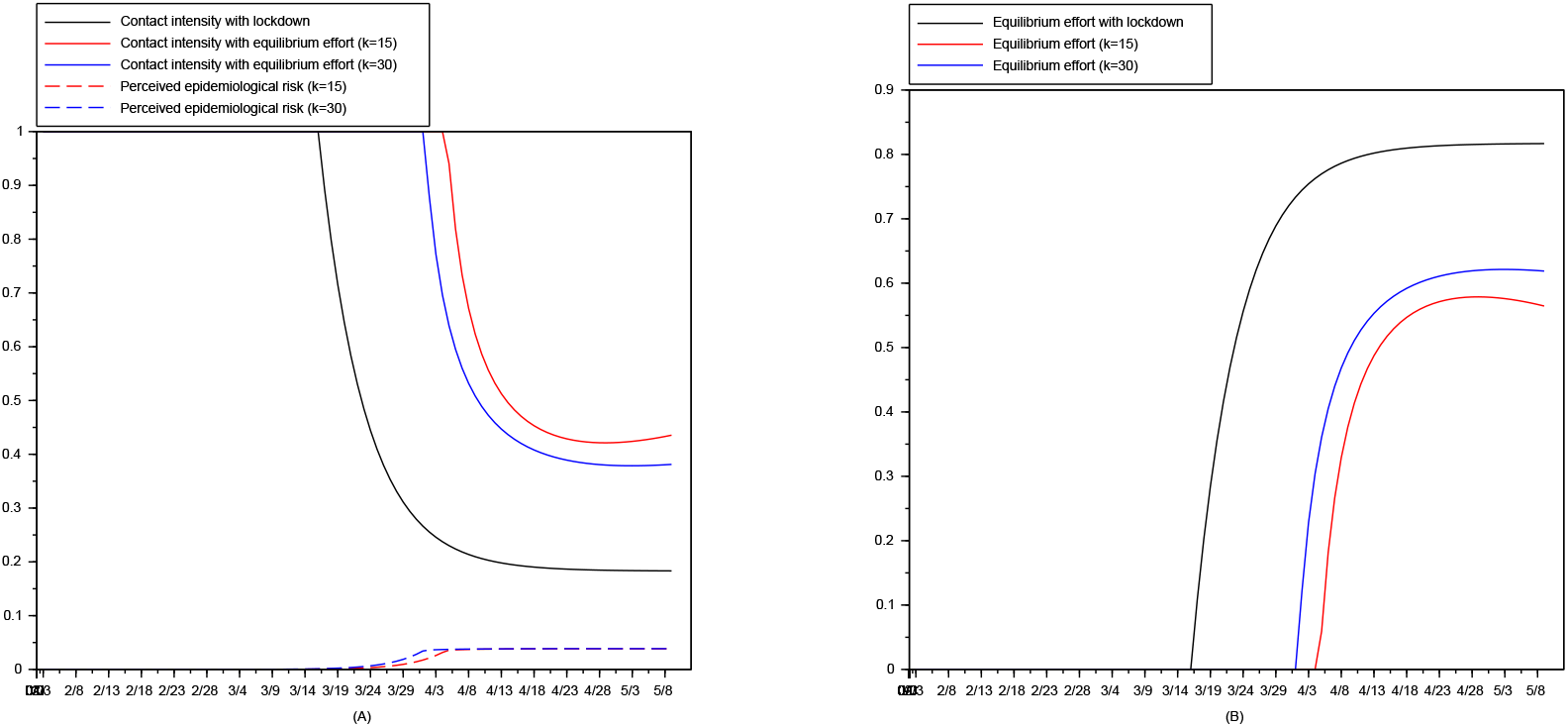
Contact intensity and equilibrium effort under various scenarios. Contact Intensity (A). Equilibrium effort (B).

With *k* = 15, contact intensity slightly increases at the end of lockdown: the number of reported cases plummets thanks to the lockdown and individual do less effort. This scenario fits survey data from Datacovid / Ipsos that indicates a weakening of distancing efforts beginning mid-April [32]. The average (self-reported) number of close contacts increased to 7.1 between May 26 and May 31, compared to 5.8 the previous week and 4.5 two weeks before [33] (data available at www.datacovid.org). We cannot directly measure the perception parameter *k*, yet *k* = 15 appears a better fit.

### Avoided deaths under lockdown and equilibrium effort

Fig. 2 contrasts the cumulative number of deaths under various scenarios. Fig. 2A shows the number of deaths in the actual situation (lockdown beginning on March 17), in the business-as-usual situation (if people had behaved as before the epidemics outbreak), and in the equilibrium self-protection model with different perceptions of disease prevalence (*k* = 15 and *k* = 30). Considering the dynamics of the number of deaths for both *k* = 15 and *k* = 30 is important given the uncertainty about the number of asymptomatic individuals and the emotional weight put on the salient COVID-19 risk.

**Figure 2:**
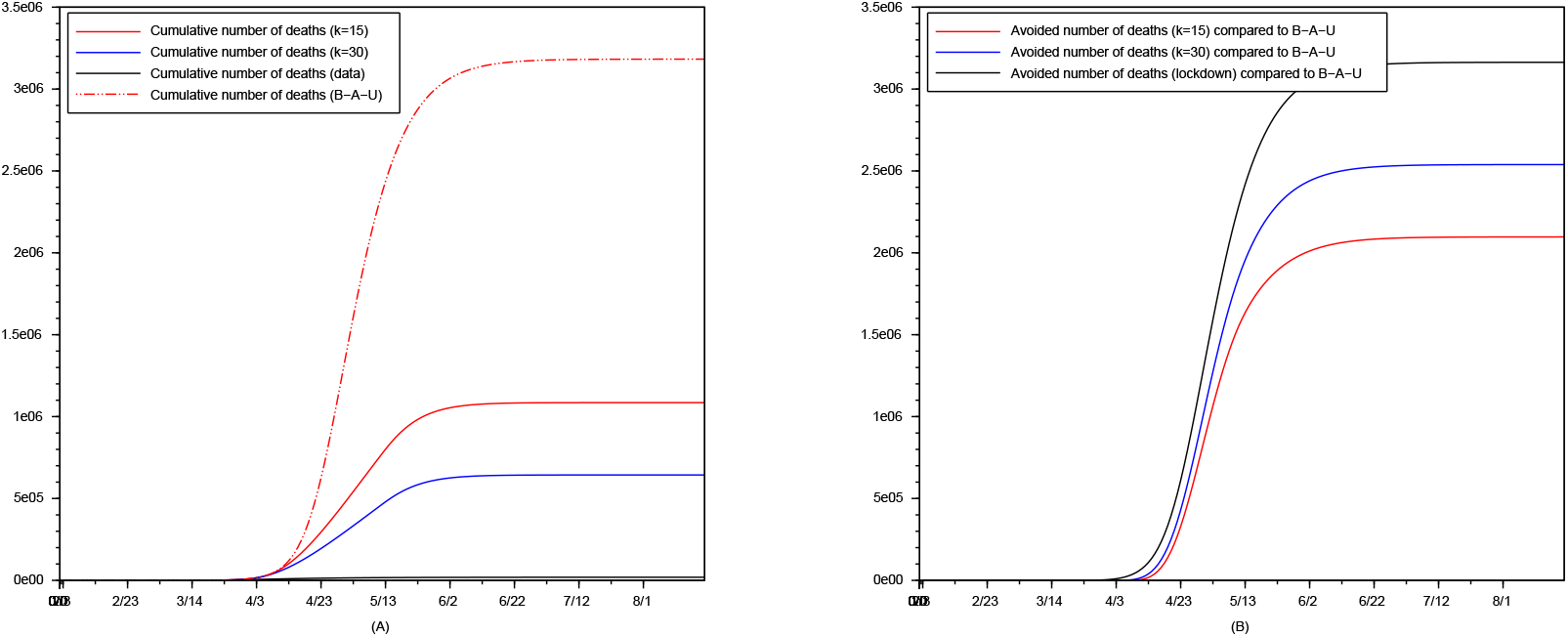
The impact of lockdown. Cumulative number of deaths under various scenarios (A). Avoided deaths compared to business-as-usual (B).

To better represent the number of deaths avoided with lockdown and with equilibrium effort, Fig. 2B graphs the difference between the cumulative number of deaths in the business-as-usual case and the other situations.

Compared with business-as-usual, the equilibrium effort with resp. *k* = 15 and *k* = 30 divides the peak number of deaths by resp. 6 and 10. Neglecting effort thus leads to an extremely large overestimation of casualties. Compared to lockdown as actually implemented, however, the equilibrium effort with resp. *k* = 15 and *k* = 30 would have lead to resp. nearly 75% and 57% more deaths. Lockdown has thus reduced the number of deaths by at least one third compared to effort alone in our simulations.

### Gradually lifting restrictions to prevent immediate rebounds

After a long lockdown period, prevalence is very low and lifting restrictions is especially attractive. Our SLIAR behavioral model however shows that a sudden lift of all restrictions can lead to a rebound, exactly – and paradoxically – because prevalence is low, so that equilibrium effort plummets. French data provides interesting evidence, because many restrictions remained imposed after full lockdown ended (May 11). An *‘intermediate lockdown’* has been instituted with strongly recommended remote work, compulsory mask wearing and priority rules for essential workers in public transportation, very limited school reopening and a 100-km limit on travel. We compute equilibrium effort and contrast data in this intermediate, gradual, stage with a counterfactual based on suddenly ending lockdown.

In the intermediate stage, [*t*_2_, *t*_3_] (May 11 – June 17), we assume that some proportion *x* of the population behaves as under full lockdown (not attending schools, working remotely,..), which corresponds to contact intensity equals to *a* = 0.2, while the remainder chooses her optimal effort, which corresponds to contact intensity *φ*^*\**^(*L*). The contact intensity function for the population used for [*t*_1_, *t*_3_] is *φ*(*t*) = *φ*_*IL*_(*t*), defined as follows.

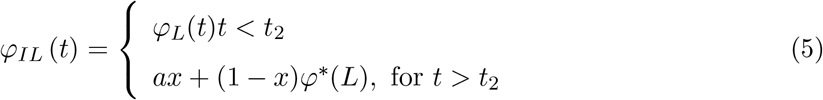

Prevalence is so low after 9 weeks of full lockdown that individual best responses lead to a null effort, as under business-as-usual, for both *k* = 15 and *k* = 30 (and any value in between): *ε*^*\**^ = 0 independently of *k*. Given this equilibrium effort, we fit the data with least-squares to estimate proportion *x*, and obtain that the degree of lockdown corresponds to *x*^*\**^ = 91.85633% (that is, an average contact intensity of 0.251, instead of 0.2 under full lockdown).

### Intermediate lockdown prevents exponential rebound

Epidemic dynamics under this continued ‘intermediate’ lockdown can be compared to two counterfactual scenarios, that is what would have happened if lockdown would had been fully maintained or on the contrary if lockdown had been fully removed, and one had solely relied on individual distancing choices.

We simulate System 1 together with contact intensity function *φ* defined by System 5, with *x* = 1, which corresponds to full lockdown, with *x* = 0, which corresponds to full removed of lockdown, and compare it to *x*^*\**^ = 91.85633% which is fitted using the data. We find that the epidemic dynamics would have exhibited *an exponential growth* in the number of cases, under both scenarios *k* = 15 and *k* = 30, Fig. 3. This exponential shape is reminiscent of the one obtained with traditional business-as-usual simulations. And indeed, we have seen that prevalence is so low at the end of the full lockdown period, that equilibrium effort is set at its lowest possible level, its lower bound 0, as in business-as-usual (yielding a contact intensity of 1).

**Figure 3:**
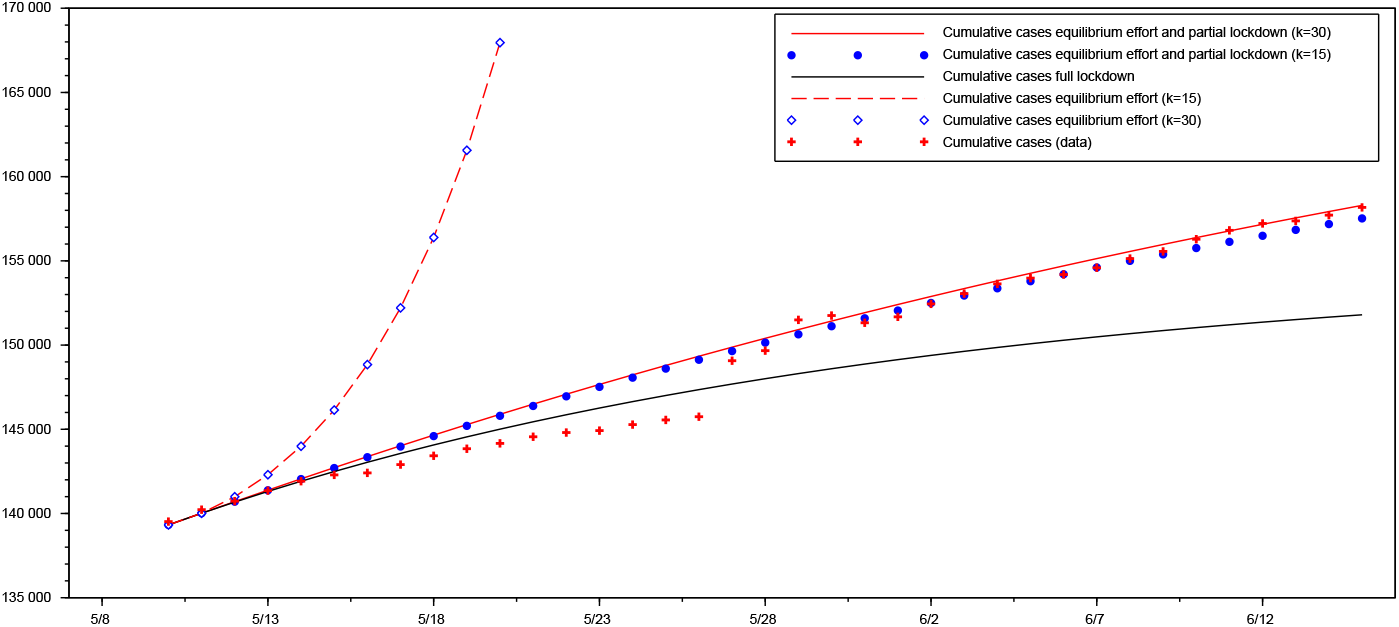
Cumulative cases in the intermediate lockdown period. Counterfactual (full lockdown), (effort under no restrictions), simulation (effort under intermediate lockdown fitted at 91.85633%), data.

In contrast, actual data shows that the number of cases increases, but in a controlled way. Intermediate lockdown thus appears effective at curbing the COVID-19 dynamics and avoiding a quick rebound.

A striking corollary is that under *very low prevalence* (as after full lockdown), beliefs and attitudes towards the infection risk play no role in the dynamics of the epidemic: Effort is the same (null), and the curves in Fig. 3 are indistinguishable for *k* = 15 and *k* = 30.

## Discussion

Equilibrium effort continuously adjusts to the number of reported cases, creating complex epidemic dynamics. We use our modelling to also analyze the interplay between equilibrium effort and various public policies after all restrictions are lifted (*t*_3_ = June 17). Our simulations are not meant to be exact predictions, given large remaining uncertainties on COVID-19 and on alternative tools (e.g., testing and tracing) not modeled here. We highlight general patters and complex interactions, between individual maximization, epidemic prevalence and policy effectiveness.

To fit the data, we assume that for *t ∈*]*t*_2_, *t*_3_[(intermediate period), *φ*(*t*) = *φ*_*IL*_(*t*). The subsequent transmission rate *φ*(*t*) for *t > t*_3_, is determined by our behavioral assumptions, with adjustments to represent specific policies.

### Distancing effort, perceptions and public salience

#### Effort after lockdown is fully removed

A main result of our SLIAR behavioral model is that individuals do not maintain a constant effort level at all times (Fig. 4). They never choose the maximum possible effort. Their effort is not proportional to reported cases either. Individuals adapt their self-protection effort in such a way that their perceived epidemiological risk remains at a (slightly downward-sloping) plateau. They do nearly no self-protection at the end of the lockdown period, since the number of reported cases is then very small. Their effort increases during a rebound, but decreases again when it is sufficient to maintain the epidemics at a low enough level. The epidemics lasts longer but has a much lower prevalence than if one neglected efforts (as in the business-as-usual scenario).

**Figure 4:**
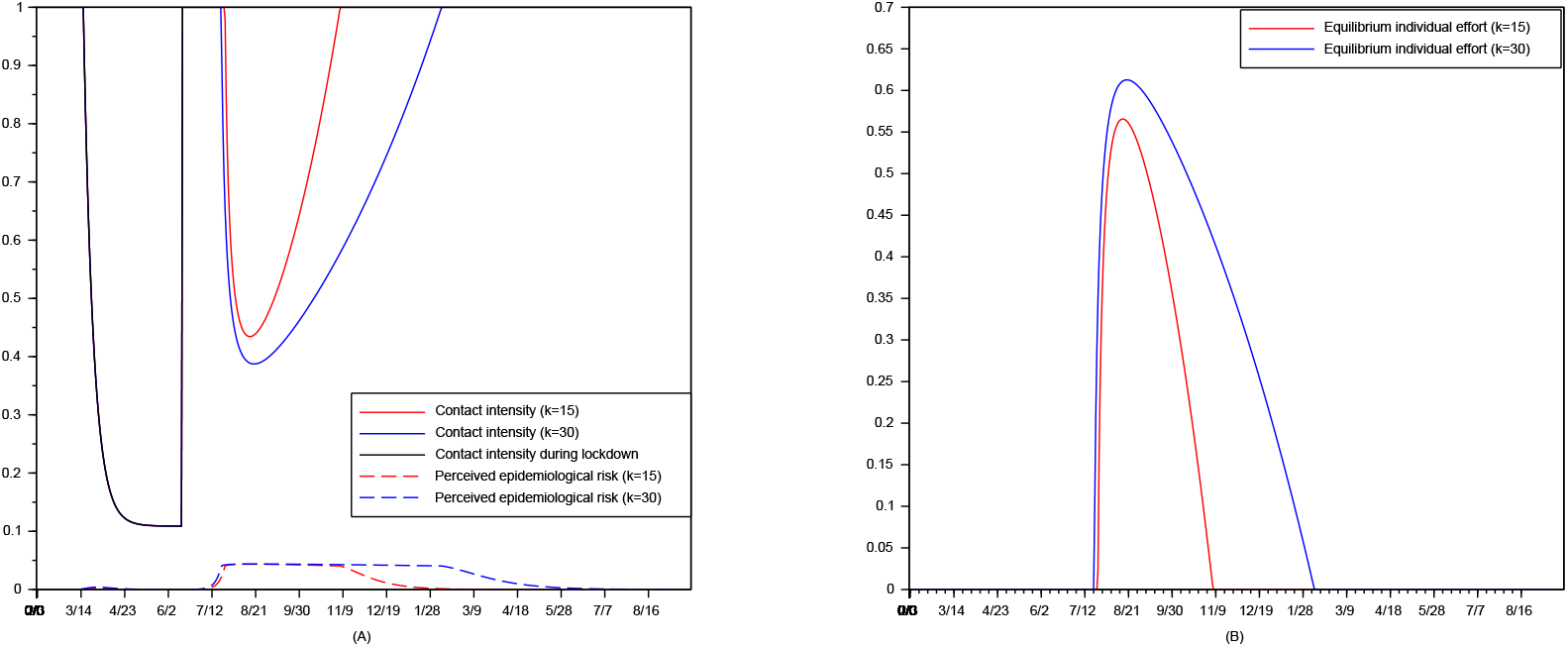
The impact of perceptions. Efforts and prevalence. Contact Intensity (A). Equilibrium effort (B).

#### Effort and perceptions

Individual beliefs about the *severity* of the disease (the utility loss when being ill, as measured by parameter *λ*) do not much affect the epidemic dynamics (results available from the authors upon request). Conversely, individual beliefs about, and attention paid to, the *infection risk*, as measured by parameter *k*, play a major role (Fig. 4). Moving from *k* = 15 to *k* = 30 increases equilibrium effort after lockdown, and effort is positive for a longer period. The epidemics has a flatter and longer plateau, and lasts about three months longer.

Thanks to this higher effort, the peak number of reported cases is divided by about 2 (Fig. 5), even though the impact on cumulative deaths is much weaker.

**Figure 5:**
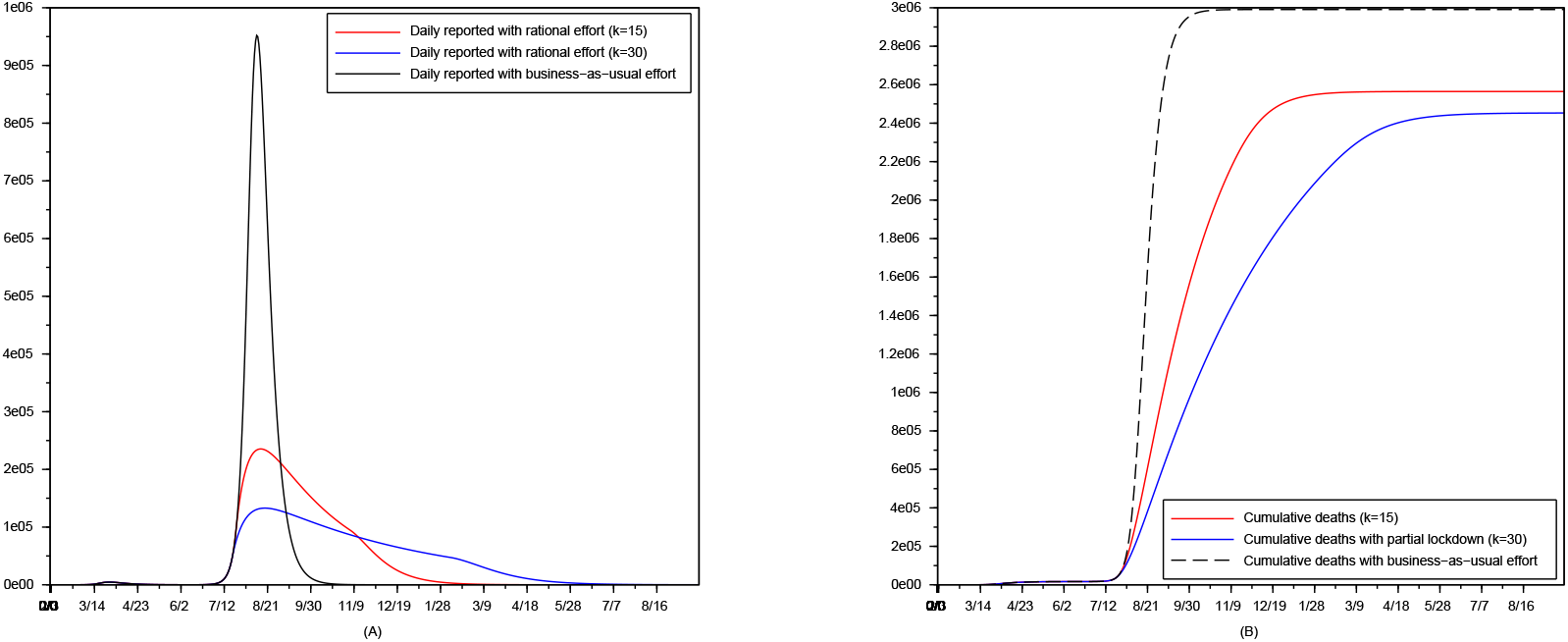
The impact of perceptions. Cases and casualties. Reported cases (A). Cumulative deaths (B).

#### Public salience

Survey data in [34] report that the main drivers of Japanese’s precautionary efforts are the highly media-covered infection aboard the Diamond Princess cruise ship (February 2020), and governmental information, both corresponding to an increase in *k*. A potentially effective public policy consists in exacerbating public awareness about the disease, with media intervention and legal measures that draw attention to the infection risk. Variations in *k* can be used to represent the impact of media coverage and public discourse, as well as the expressive power of legal measures. This is a double-sword tool: Full lockdown removal may be interpreted as a signal of a low infection risk, leading to a drop in *k*.

Importantly, effort can increase when perception of infection risk increases (as measured by *k*) despite the lower epidemics prevalence associated with this higher effort. This is a specific advantage of the communication policy.

### Public policies partly crowd out individual efforts

We use our model to simulate the impact of various public policies on the COVID-19 epidemics, given individuals’ reaction to reported cases. The effectiveness of policies indeed strongly depends on the side impact they have on equilibrium self-protection efforts.

#### Partial lockdown

Imposing partial lockdown (for instance for vulnerable population or employees who have the ability to work remotely) helps reduce the cumulative number of deaths. Compared to 30%-lockdown, a 60% lockdown further displaces the curve of reported cases, and lowers the peak level. This comes at the expense of a duration of the epidemic increased by one month, as immunity grows more slowly. This stricter lockdown is mostly effective at markedly reducing the number of deaths in the long run. A lockdown of 30% [resp. 60%]of the population decreases the peak number of cases by 20% [resp. 46%]. Partial lockdown delays the rebound and flattens the epidemic dynamics (Fig. 6).

**Figure 6:**
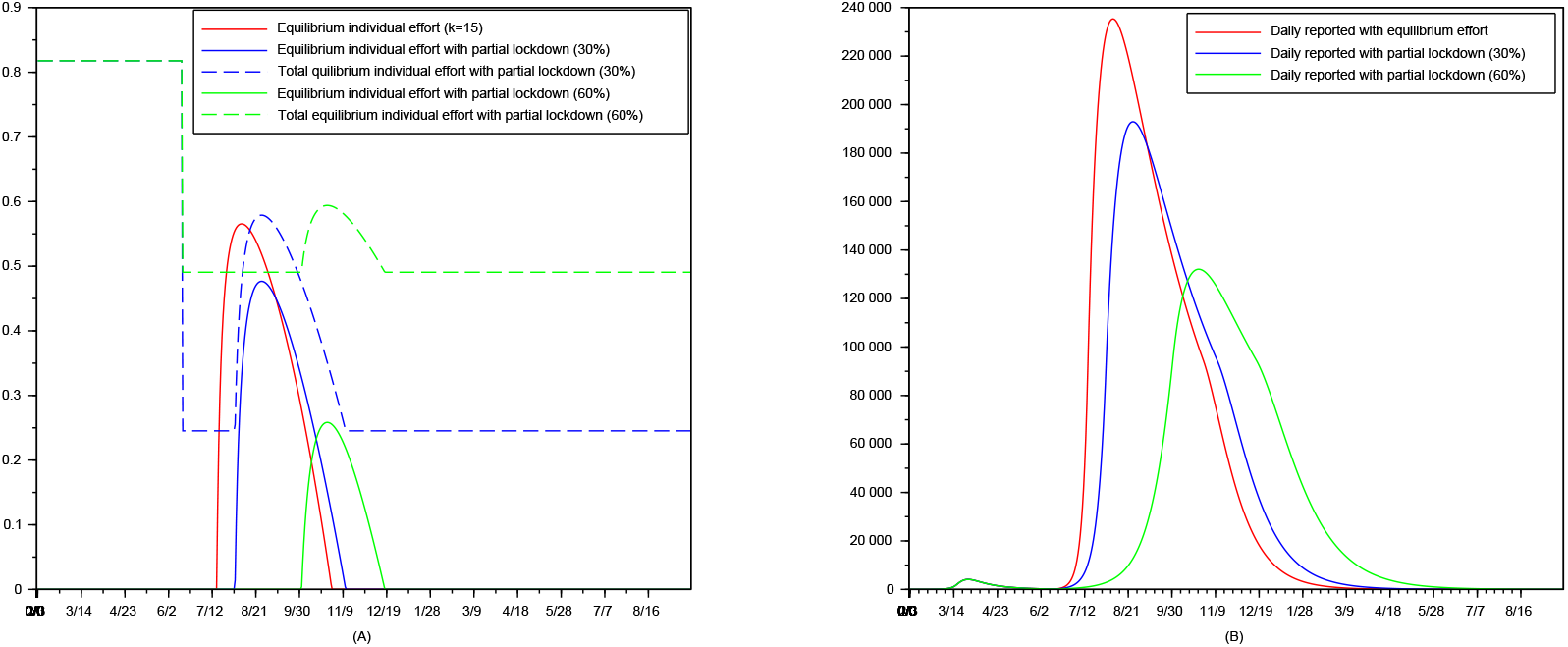
The impact of partial lockdown, *k* = 15. Physical distancing effort (A) and epidemics dynamics (B). Horizontal (dotted lines) segments correspond to a null freely chosen effort, averaged with compulsory effort for a fraction of the population.

Partial lockdown however reduces not only infection prevalence, but also effort. Because there are fewer new cases than in the absence of any lockdown, effort decreases in the percentage of the population that remains under lockdown. It plummets with a 60% lockdown. A large part of the potential benefits of partial lockdown thus disappears due to this downward adjustment of effort. This is even more so when effort would have been higher anyway due to a higher perception of the risk (Fig. 7, *k* = 30).

**Figure 7:**
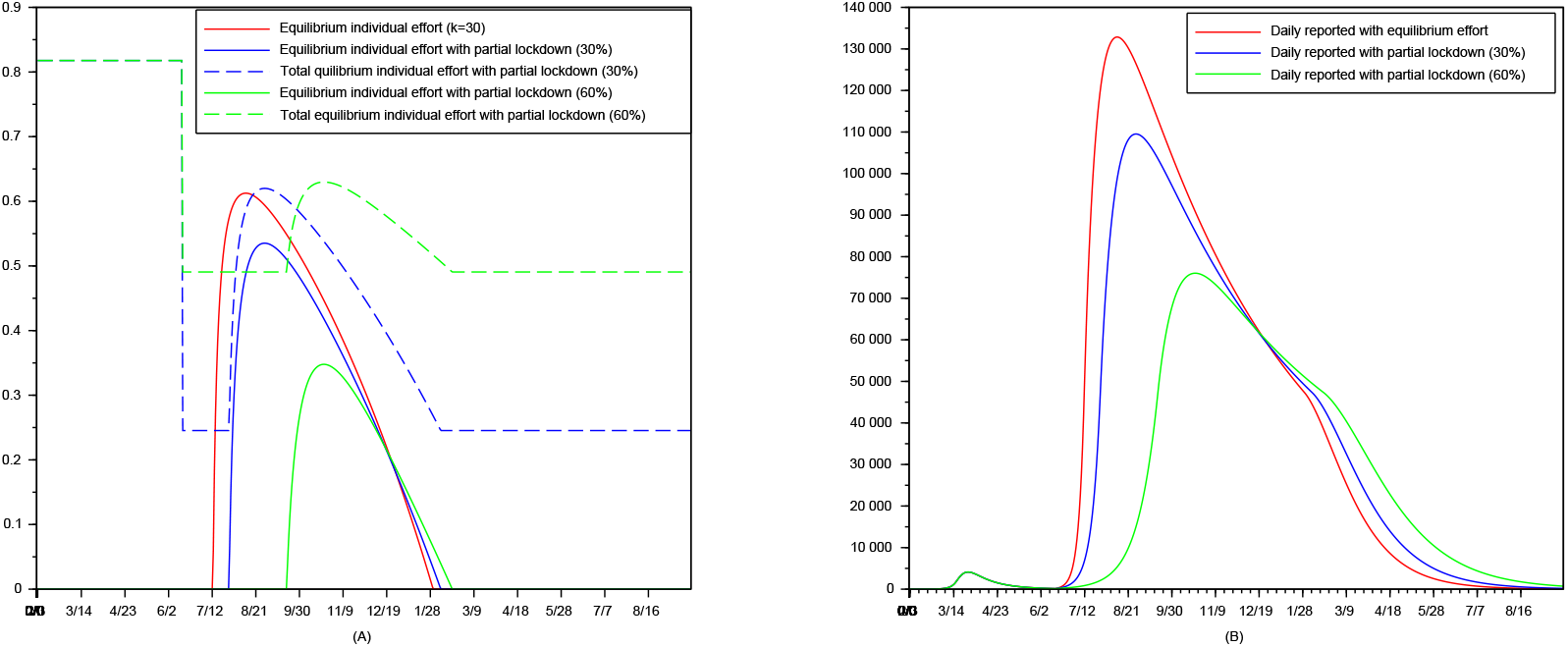
The impact of partial lockdown, *k* = 30. Physical distancing effort (A) and epidemics dynamics (B).

This crowding-out effect is not sufficiently large in our context to make lockdown inefficient in terms of epidemics dynamics. It questions however the desirability of such partial lockdown given its economic and psychological costs. Indeed, the benefit of partial lockdown is due to the fact that it maintains a low average contact intensity despite the decrease in prevalence. While this helps flatten the epidemic spread compared with freely chosen effort, it comes at the cost of high levels of distancing for long periods and low prevalence.

#### Compulsory mask wearing in public places

Wearing a mask protects others from transmission [35] but has little direct effect on self-protection. We assume that individuals are aware of that. In an economic equilibrium, absent altruism and social norms, they would not wear masks if it involves some disutility or cost. Compulsory mask wearing for some proportion *m* of activities (for instance in public transports) however reduces the global transmission risk: Perceived risk is now

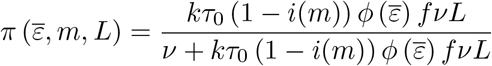

where *i*(*m*) is the impact of doing the legally imposed effort *m* of wearing a mask.

We use *i*(*m*) = 1 *− e*^0.1*m*^ and *m* = 2, so that 1 *− i*(*m*) is 18%. This corresponds to masks blocking 82% of the virus transmission in the population. Given that masks effectiveness vary widely, that individuals may not properly wear and care for their masks and that some may not comply [36], this figure yields *upper bounds* on the benefits of compulsory mask wearing.

Mask wearing partly blocks virus transmission, so individuals observe that there are fewer reported cases, and they choose to less reduce their contacts in equilibrium. This counteracts the protective effect of the mask for others. Its overall effect is not straightforward.

In our calibration, with an 82% reduction in virus transmission, the total effect of compulsory mask wearing remains positive (Fig. 8) but its impact is quite small: The reported cases and cumulative deaths are *nearly indistinguishable* with and without masks (Fig. 9). The overall dynamics of the reported number of cases is nearly unchanged, and the number of deaths weakly decreases. The lack of impact of masks on epidemic dynamics comes from a crowding out effect: Compulsory mask wearing reduces the peak distancing effort level by 1/6. This negative impact is compensated for in the transmission process by the effectiveness of the mask at protecting others (*i*(*m*)). If mask wearing happens to be less effective than the 82% we use, its overall effect can be negative on transmission. This holds even though we assume that individuals do not believe that the mask protects them.

**Figure 8:**
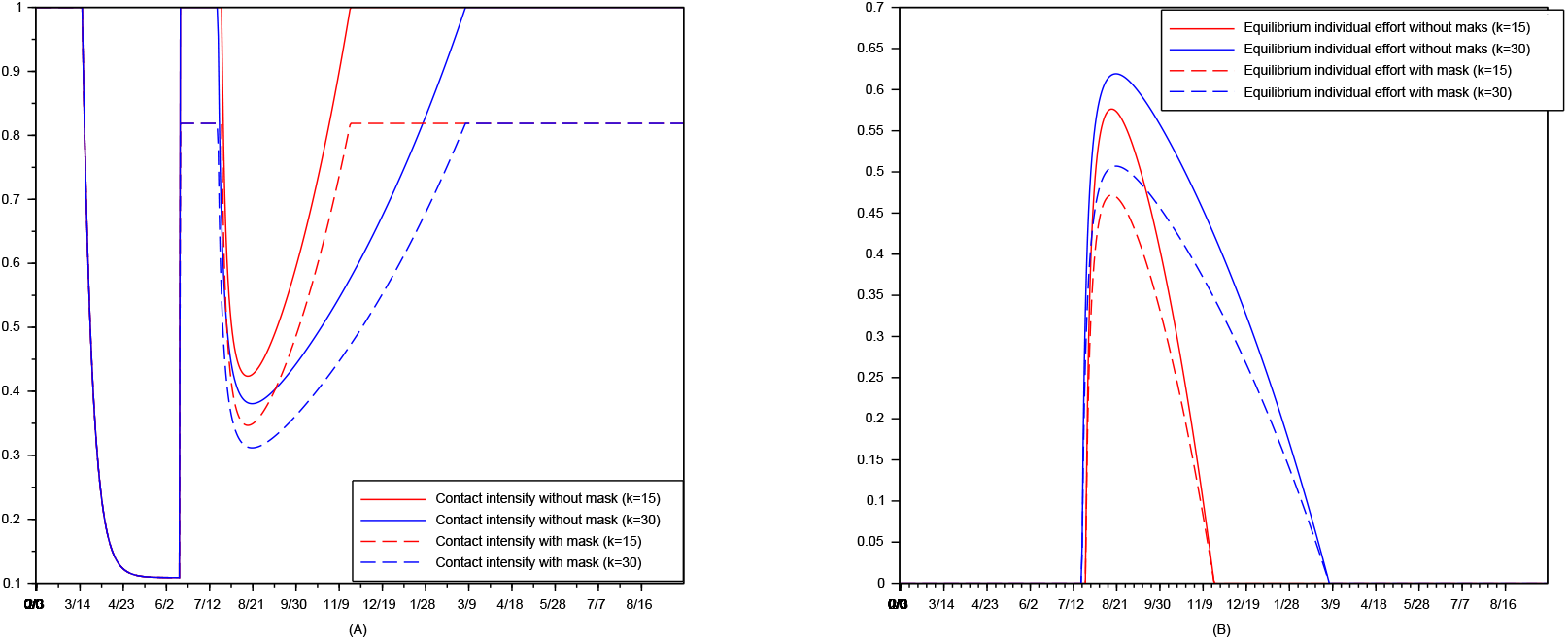
The impact of compulsory mask wearing. Contact Intensity (A). Equilibrium effort (B).

**Figure 9:**
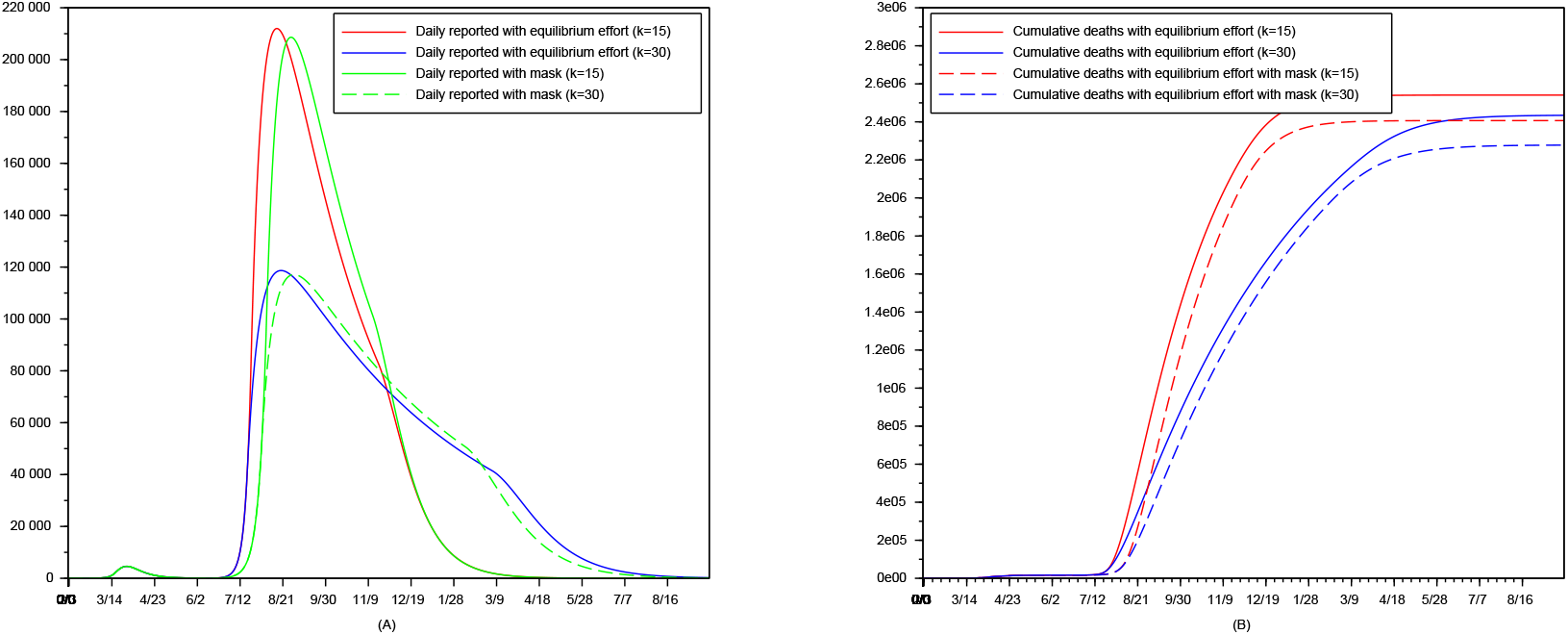
The impact of compulsory mask wearing. Reported cases (A). Cumulative deaths (B).

Because masks help individuals increase contacts intensity without triggering an epidemics surge, they improve social welfare. However it is striking that the impact of compulsory mask wearing lies in the reduction of the economic and well-being costs from distancing, but *not* in a noticeable reduction in epidemic prevalence.

## Conclusion

Our results provide insight on public debates: Ignoring distancing choices likely leads to a strong overestimation of the number of deaths avoided thanks to lockdown; but lockdown saves at least one third more lives than freely chosen efforts. Policies post-lockdown crowd out self-protection efforts so that their overall effectiveness is limited. Mask wearing helps reinstate contacts, but has nearly no impact on the epidemic. Communication appears desirable as it can increase effort for a given reported prevalence.

## Data Availability

All data use is publicly available from the French national health agency. https://www.data.gouv.fr/en/datasets/donnees-hospitalieres-relatives-a-lepidemie-de-covid-19-en-france/

https://dashboard.covid19.data.gouv.fr

https://www.data.gouv.fr/en/datasets/donnees-hospitalieres-relatives-a-lepidemie-de-covid-19-en-france/

